# Broad-spectrum of non-serious adverse events following COVID-19 vaccination: A population-based cohort study in Seoul, South Korea

**DOI:** 10.1101/2023.11.15.23298566

**Authors:** Jee Hyun Suh, Hong Jin Kim, Min-Ho Kim, Myeong Geun Choi, Eun Mi Chun

## Abstract

Much of the current literature on the adverse effects occurring after the coronavirus disease-2019 (COVID-19) vaccination focused on serious adverse events (AEs). Consequently, the non-serious AEs have yet to be comprehensively elucidated. This study aims to investigate the incidence rate and risk of non-serious AEs including gynecological, hematological, dermatological, ophthalmological, otologic, and dental problems following the COVID-19 vaccination. We conducted a population-based cohort study was conducted with the National Health Insurance Service (NHIS) database in Seoul, South Korea. The cumulative incidence rate (cIR) per 10,000 population, Odds ratio, and Hazard ratio (HR) with a 95% Confidence Interval (CI) were measured to assess the non-serious AEs, as reported by the Vaccine Adverse Event Reporting Center, after COVID-19 vaccination. The cIR of non-serious AEs for three months was significantly higher in vaccinated subjects than in non-vaccinated subjects, except for endometriosis. The vaccination significantly increased the risks of all the non-serious AEs except for visual impairment. The risk of inner ear disease showed the highest HRs (HR [95% CI] = 2.37 [2.15-2.60]) among the non-serious AEs following COVID-19 vaccination. Among the vaccinated subjects, heterologous vaccination was associated with the increased risk of most of the non-serious AEs. The three-month risks of incidental non-serious AEs are substantially higher in the COVID-19 vaccinated subjects than in non-vaccinated controls. Our findings suggested that vaccinated subjects with predisposition are potentially vulnerable to the occurrence of broad-spectrum AEs although the COVID-19 vaccines may not be serious.

## Introduction

Coronavirus disease-2019 (COVID-19), as a global challenge for health and socioeconomic issues, demonstrated a subsequent rise in morbidity and mortality in the early stages of the pandemic compared to other viral infections [1]. With the rapid development of vaccines in response to the unprecedented COVID-19 pandemic, there has been a contribution to the reduction of the severity and fatality rates. Subsequently, several types of vaccines after approval of the AZD1222 vaccine have been released to prevent COVID-19 infection [2–5]. However, a wide range of adverse events (AEs), not previously reported in conventional vaccines, have been observed post-vaccination [6–8].

As every country has done, South Korea initiated the COVID-19 vaccination program at care facilities and subsequently expanded its coverage to encompass the entire nation’s population. About 80% of the population in South Korea was vaccinated within a year, which contributed to a significant decrease in COVID-19 infection [9]. Meanwhile, there is increasing evidence that many vaccinated populations could experience several unexpected complications, such as immune-related AEs (AEs) [6–8, 10, 11]. Many post-vaccination AEs are believed to originate from an immune response characterized by an inflammatory cytokine storm that causes irreversible damage to the cardiovascular, cerebrovascular, and respiratory systems [12, 13].

Much of the current literature on the side effects occurring after the COVID-19 vaccination focused on serious AEs such as cardiovascular complications. Consequently, the non-serious AEs have yet to be comprehensively elucidated [11, 14–20]. Therefore, this study aims to assess the non-serious AEs after COVID-19 vaccination from the National Health Insurance Service (NHIS) database in Seoul, South Korea.

## Methods

### Data source

This from the Korean NHIS database on 1, January 2021 enrolled randomly extracted 50% of individuals residing in Seoul, South Korea, as a representative sample. We randomly selected 50% of the residents living in Seoul as of January 1, 2021. The International Classification of Diseases, 10th revision (ICD-10), was adopted by the NHIS to classify disease diagnoses. The data included the primary diagnosis, secondary diagnosis, and dates of hospital visits. This population-based cohort study was conducted by the Strengthening the Reporting of Observational Studies in Epidemiology (STROBE) guidelines [21].

### Study population

A total of 4,348,412 individuals living in Seoul, constituting 50% of the population, were included and investigated as of January 1, 2021. Individuals aged under 20 years were excluded, leaving 4,203,887 individuals for analysis. In this study, only individuals who had received two doses of COVID-19 vaccine were included in the vaccinated group. In this cohort study, the index date, which is the date on which individuals started participating in the study, was set differently for the vaccinated and non-vaccinated groups. For the vaccinated group, the index date was set as the date of the second vaccine dose administered before October 1, 2021. On the other hand, for the non-vaccinated group, the index date was set as October 1, 2021. The vaccinated group included 3,839,014 individuals, whereas the non-vaccinated group included 364,873 individuals. Individuals who received a dose of vaccine before January 1, 2021, and did not receive a second dose between January 1, 2021, and October 1, 2021, were excluded. The vaccinated group included 2,154,389 individuals, whereas the non-vaccinated group included 350,953 individuals.

The non-serious AEs included gynecological (endometriosis, and menstrual disorders [polymenorrhagia, menorrhagia, abnormal cycle length, oligomenorrhea, and amenorrhea]), hematological (bruises confined to non-tender and yellow-colored on especially extremities), dermatological (herpes zoster, alopecia, and warts), ophthalmological (visual impairment, and glaucoma), otological (tinnitus, inner ear, middle ear, and outer ear disease), and dental problems (periodontal disease) as reported by the Vaccine Adverse Event Reporting Center. To investigate the causal relationship between vaccine administration and AEs, the diagnostic records for a year before the index date were traced. Individuals with any target disease as a primary or secondary diagnosis during this period were excluded from the study. The occurrence of the target disease was defined as receiving a primary diagnosis of the disease from the day after the index date.

### Outcome measurements

The primary outcome measure was cumulative incidence rates (cIRs) of AEs per 10,000 population between the vaccinated and non-vaccinated subjects. The cIRs of the AEs were measured at one week, two weeks, one month, and three months. The secondary outcome measures were the odd ratios (ORs) and hazard ratios (HRs) of AEs. Furthermore, subgroup analyses were also conducted based on gender, the number of COVID-19 vaccine doses, the vaccine type (mRNA vaccine, cDNA vaccine, and heterologous vaccination), health insurance level, presence of diabetes mellitus (DM), hypertension (HTN), hyperlipidemia, and chronic obstructive pulmonary disease (COPD).

Age, gender, insurance level, Charlson’s comorbidity index (CCI), presence of DM, HTN, hyperlipidemia, and COPD, and prior COVID-19 infection history were extracted using their ICD-19 codes, which were suggested by Sundararajan et al. The presence of comorbid diseases (i.e. DM, HTN, hyperlipidemia, and COPD), items in CCI, and the prior COVID-19 infection history was determined as a primary or secondary diagnosis 2 or more times within 1 year before the index date [22]. The NHI premium was used as a proxy measure of income because it is proportional to monthly income, including earnings and capital gains. The income quantiles of the enrolled individuals were categorized into three groups (low-, middle- and high-income groups in medical aid enrollees and the 0–33, 34–66, and 67–100 centiles of NHI enrollees).

### Statistical analysis

Statistical analysis was performed using SAS Enterprise Guide (version 8.3., SAS Institute, Cary, NC, USA). A normal distribution was confirmed with the Kolmogorov–Smirnov test. Baseline patient characteristics and comorbidities were reported as means ± standard deviation for continuous variables and frequency (percentage, %) for categorical variables. Student’s t-test was performed for continuous variables, and the chi-square test or immune-mediated adverse events associated with COVID-19 vaccination were assessed using Student’s t-test for continuous variables, and the chi-square test or Fisher’s exact test for categorical variables. The cIR was calculated per 10,000 populations. To identify the association between COVID-19 vaccination and AEs, a multiple logistic regression model was used for ORs, corresponding to 95% CIs. Cox proportional hazards regression was used to estimate the HRs and 95% CIs. Two-sided *P* values of 0.05 or less were considered to indicate statistical significance

## Results

### The participants’ characteristics

In total, 1,748,136 subjects were included in this study. Among them, 289,579 (16.57%) had not received the COVID-19 vaccine (i.e. non-vaccinated subjects), whereas 1,458,557 (83.43%) were vaccinated against COVID-19 (i.e. vaccinated subjects) (Fig. 1). The baseline characteristics of the vaccinated and non-vaccinated groups are shown in Table 1.

**Fig. 1.**
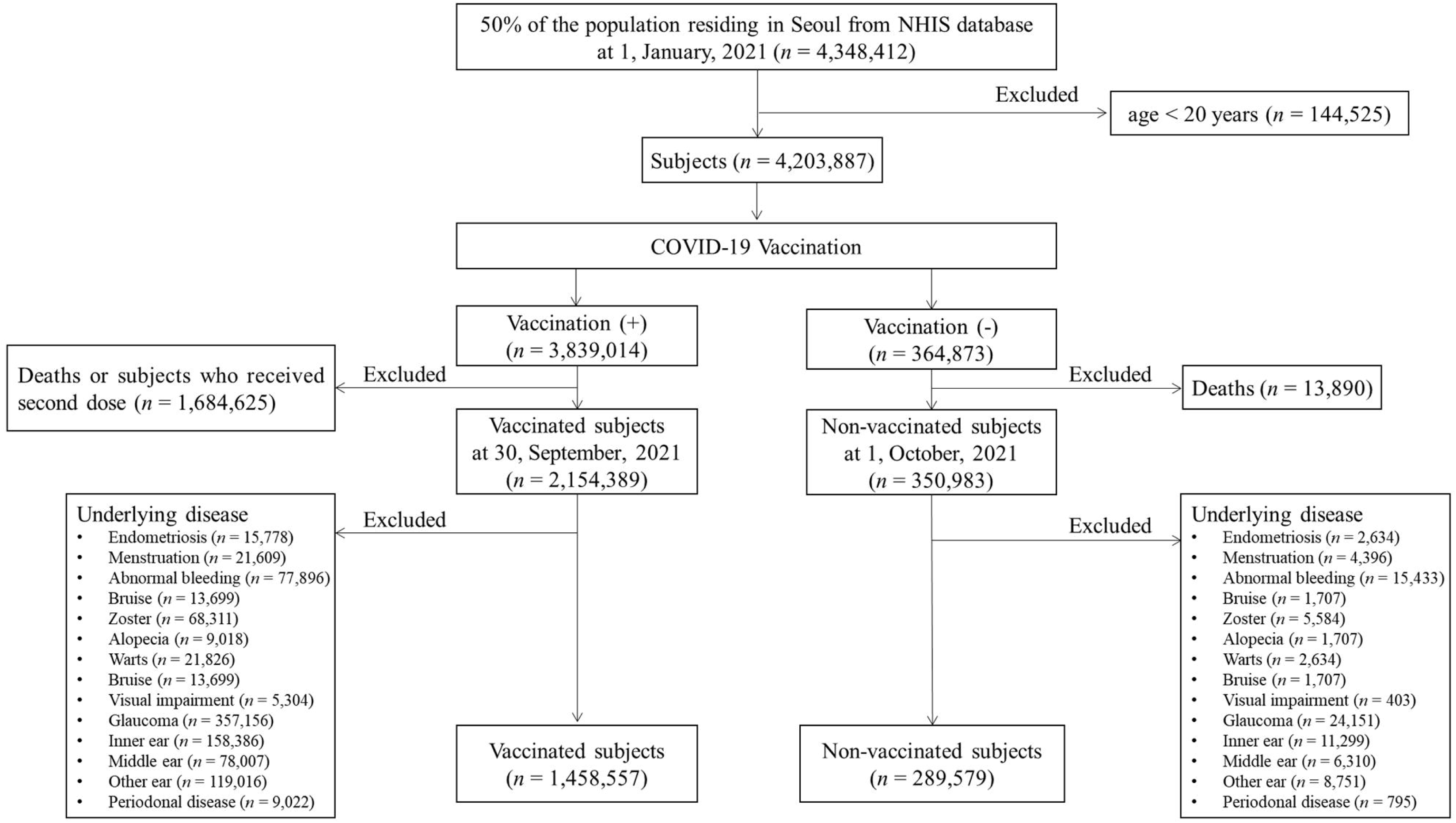
Flowchart of this study

**Table 1.**
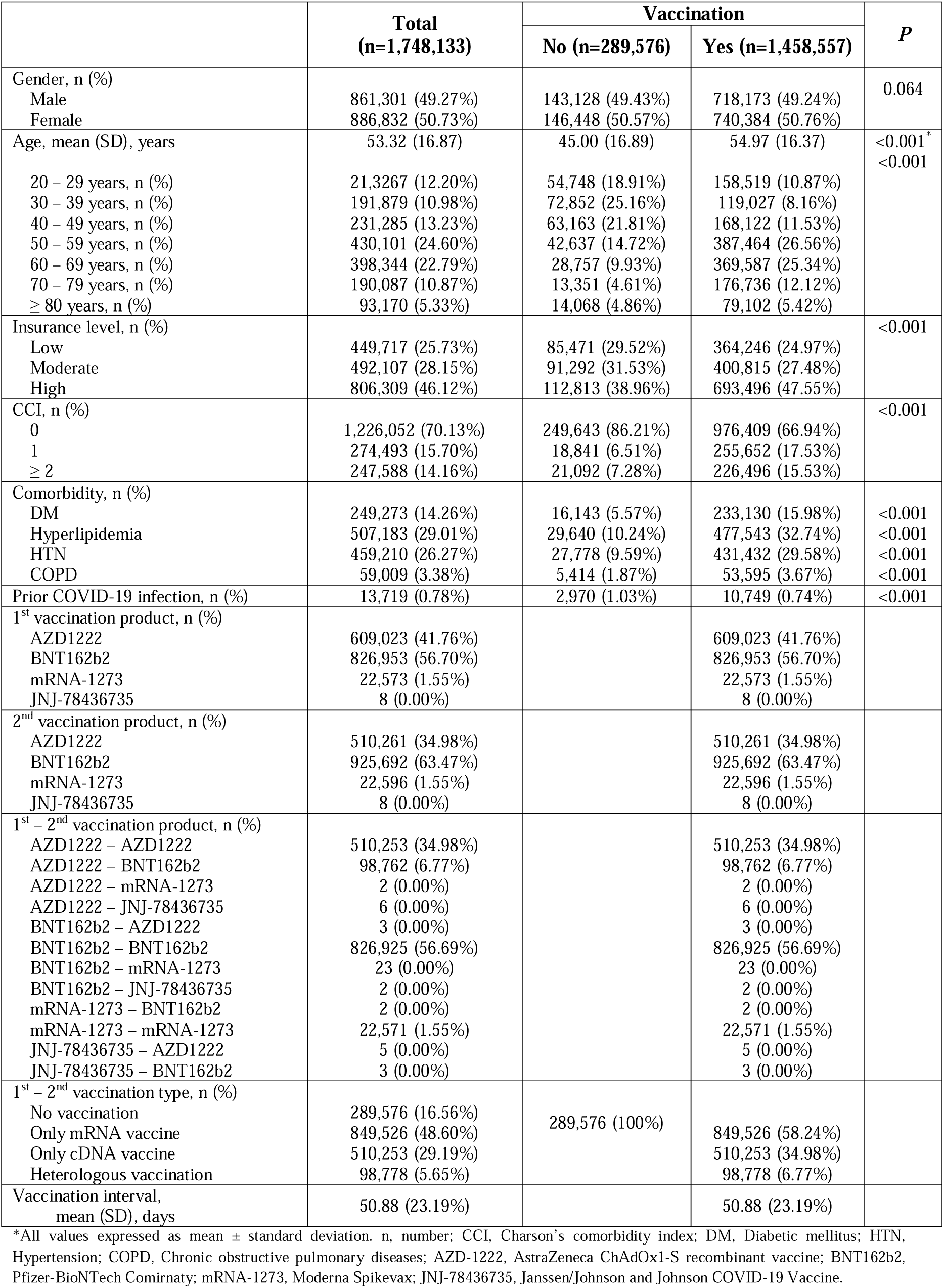
Baseline characteristics of the patients stratified by COVID-19 vaccination in South Korea.

### The cIRs per 10,000 of the non-serious AEs following the COVID-19 vaccination

Among the non-serious AEs in this study, the cIRs at three months following COVID-19 vaccination were higher in vaccinated subjects than in non-vaccinated subjects, except for endometriosis. The highest cIR of the non-serious AEs in vaccinated subjects was observed in other ear diseases (cIR, 51.78%; 95% CI, 50.61-52.94) followed by inner ear diseases (cIR, 47.10%; 95% CI, 45.99-48.21), herpes zoster (cIR, 45.08%; 95% CI, 43.99-46.17), menstrual disorders (cIR, 44.43%; 95% CI, 43.35-45.51), and glaucoma (cIR, 39.42%; 95% CI, 38.40-40.43). Among the non-serious AEs, 50% exhibited a significant difference in cIRs one week post-vaccination. Menstrual disorders and visual impairments were noted from one month, whereas alopecia, warts, and periodontal disease were observed from two weeks onwards (Table 2).

**Table 2.** Cumulative incidence rate (cIR) of non-serious adverse events following COVID-19 vaccination.

| Disease | Vaccination | Total number | One week |  |  |  | Two weeks |  |  |  | One month |  |  |  | Three months |  |  |  |
| --- | --- | --- | --- | --- | --- | --- | --- | --- | --- | --- | --- | --- | --- | --- | --- | --- | --- | --- |
|  |  |  | event | IR | 95% CI | P | event | cIR | 95% CI | P | event | cIR | 95% CI | P | event | cIR | 95% CI | P |
| Endometriosis | No | 289,576 | 4 | 0.14 | 0.00-0.27 | 0.389 | 13 | 0.45 | 0.20-0.69 | 0.877 | 35 | 1.21 | 0.81-1.61 | 0.704 | 99 | 3.42 | 2.75-4.09 | 0.15 |
|  | Yes | 1,458,557 | 34 | 0.23 | 0.15-0.31 |  | 63 | 0.43 | 0.33-0.54 |  | 165 | 1.13 | 0.96-1.30 |  | 584 | 4 | 3.68-4.33 |  |
| Menstrual disorder | No | 289,576 | 82 | 2.83 | 2.22-3.44 | 0.638 | 161 | 5.56 | 4.70-6.42 | 0.096 | 346 | 11.95 | 10.69-13.21 | 0.002 | 1025 | 35.4 | 33.23-37.56 | <0.001 |
|  | Yes | 1,458,557 | 442 | 3.03 | 2.75-3.31 |  | 935 | 6.41 | 6.00-6.82 |  | 2081 | 14.27 | 13.65-14.88 |  | 6481 | 44.43 | 43.35-45.51 |  |
| Bruise | No | 289,576 | 4 | 0.14 | 0.00-0.27 | <0.001 | 6 | 0.21 | 0.04-0.37 | <0.001 | 11 | 0.38 | 0.16-0.60 | <0.001 | 48 | 1.66 | 1.19-2.13 | <0.001 |
|  | Yes | 1,458,557 | 107 | 0.73 | 0.59-0.87 |  | 181 | 1.24 | 1.06-1.42 |  | 287 | 1.97 | 1.74-2.20 |  | 559 | 3.83 | 3.51-4.15 |  |
| Herpes zoster | No | 289,576 | 31 | 1.07 | 0.69-1.45 | <0.001 | 66 | 2.28 | 1.73-2.83 | <0.001 | 162 | 5.59 | 4.73-6.46 | <0.001 | 454 | 15.68 | 14.24-17.12 | <0.001 |
|  | Yes | 1,458,557 | 472 | 3.24 | 2.94-3.53 |  | 1055 | 7.23 | 6.80-7.67 |  | 2270 | 15.56 | 14.92-16.20 |  | 6575 | 45.08 | 43.99-46.17 |  |
| Alopecia | No | 289,576 | 6 | 0.21 | 0.04-0.37 | 0.101 | 10 | 0.35 | 0.13-0.56 | 0.003 | 23 | 0.79 | 0.47-1.12 | <0.001 | 80 | 2.76 | 2.16-3.37 | <0.001 |
|  | Yes | 1,458,557 | 61 | 0.42 | 0.31-0.52 |  | 122 | 0.84 | 0.69-0.98 |  | 266 | 1.82 | 1.60-2.04 |  | 766 | 5.25 | 4.88-5.62 |  |
| Warts | No | 289,576 | 20 | 0.69 | 0.39-0.99 | 0.279 | 30 | 1.04 | 0.67-1.41 | <0.001 | 78 | 2.69 | 2.10-3.29 | <0.001 | 197 | 6.8 | 5.85-7.75 | <0.001 |
|  | Yes | 1,458,557 | 135 | 0.93 | 0.77-1.08 |  | 303 | 2.08 | 1.84-2.31 |  | 658 | 4.51 | 4.17-4.86 |  | 1811 | 12.42 | 11.84-12.99 |  |
| Visual impairment | No | 289,576 | 0 | 0 | 0.00-0.00 | 1.00 | 0 | 0 | 0.00-0.00 | 0.372 | 0 | 0 | 0.00-0.00 | 0.024 | 2 | 0.07 | 0.00-0.16 | 0.008 |
|  | Yes | 1,458,557 | 5 | 0.03 | 0.00-0.06 |  | 9 | 0.06 | 0.02-0.10 |  | 23 | 0.16 | 0.09-0.22 |  | 50 | 0.34 | 0.25-0.44 |  |
| Glaucoma | No | 289,576 | 43 | 1.48 | 1.04-1.93 | <0.001 | 80 | 2.76 | 2.16-3.37 | <0.001 | 199 | 6.87 | 5.92-7.83 | <0.001 | 534 | 18.44 | 16.88-20.00 | <0.001 |
|  | Yes | 1,458,557 | 430 | 2.95 | 2.67-3.23 |  | 901 | 6.18 | 5.77-6.58 |  | 1892 | 12.97 | 12.39-13.56 |  | 5749 | 39.42 | 38.40-40.43 |  |
| Tinnitus | No | 289,576 | 11 | 0.38 | 0.16-0.60 | 0.013 | 26 | 0.9 | 0.55-1.24 | 0.003 | 54 | 1.86 | 1.37-2.36 | <0.001 | 171 | 5.91 | 5.02-6.79 | <0.001 |
|  | Yes | 1,458,557 | 119 | 0.82 | 0.67-0.96 |  | 237 | 1.62 | 1.42-1.83 |  | 534 | 3.66 | 3.35-3.97 |  | 1789 | 12.27 | 11.70-12.83 |  |
| Inner ear disease | No | 289,576 | 43 | 1.48 | 1.04-1.93 | <0.001 | 71 | 2.45 | 1.88-3.02 | <0.001 | 152 | 5.25 | 4.41-6.08 | <0.001 | 466 | 16.09 | 14.63-17.55 | <0.001 |
|  | Yes | 1,458,557 | 553 | 3.79 | 3.48-4.11 |  | 1094 | 7.5 | 7.06-7.94 |  | 2381 | 16.32 | 15.67-16.98 |  | 6870 | 47.1 | 45.99-48.21 |  |
| Middle ear disease | No | 289,576 | 14 | 0.48 | 0.23-0.74 | <0.001 | 42 | 1.45 | 1.01-1.89 | <0.001 | 93 | 3.21 | 2.56-3.86 | <0.001 | 290 | 10.01 | 8.86-11.17 | <0.001 |
|  | Yes | 1,458,557 | 218 | 1.49 | 1.30-1.69 |  | 468 | 3.21 | 2.92-3.50 |  | 1058 | 7.25 | 6.82-7.69 |  | 3343 | 22.92 | 22.14-23.70 |  |
| Other ear disease | No | 289,576 | 43 | 1.48 | 1.04-1.93 | <0.001 | 86 | 2.97 | 2.34-3.60 | <0.001 | 202 | 6.98 | 6.01-7.94 | <0.001 | 607 | 20.96 | 19.30-22.63 | <0.001 |
|  | Yes | 1,458,557 | 550 | 3.77 | 3.46-4.09 |  | 1112 | 7.62 | 7.18-8.07 |  | 2441 | 16.74 | 16.07-17.40 |  | 7552 | 51.78 | 50.61-52.94 |  |
| Periodontal disease | No | 289,576 | 5 | 0.17 | 0.02-0.32 | 1.00 | 6 | 0.21 | 0.04-0.37 | 0.044 | 14 | 0.48 | 0.23-0.74 | 0.001 | 31 | 1.07 | 0.69-1.45 | <0.001 |
|  | Yes | 1,458,557 | 30 | 0.21 | 0.13-0.28 |  | 70 | 0.48 | 0.37-0.59 |  | 160 | 1.1 | 0.93-1.27 |  | 488 | 3.35 | 3.05-3.64 |  |

Cumulative incidence rates were calculated as a rate per 10,000 individuals. IR, incidence rate; CI, confidence interval; cIR, cumulative incidence rate.

When stratified by gender, the cIRs of AEs showed a similar pattern to that of the overall population. At three months post-vaccination, menstrual disorders presented the highest cIRs in females (cIR, 87.54%; 95% CI, 85.41-89.66) followed by inner ear diseases in females (cIR, 62.82%; 95% CI, 61.02-64.62), other ear diseases in females (cIR, 55.27%; 95% CI, 53.58-56.96), and herpes zoster in females (cIR, 53.13%; 95% CI, 51.48-54.79). When stratified by vaccine type, heterologous vaccination increased the cIR of menstrual disorders to 78.96% (95% CI, 73.45-84.48). The detailed cIRs stratified by gender or vaccine type were presented in eTables 1 and 2.

### The risks of non-serious AEs following the COVID-19 vaccination

In the Cox proportional hazard model in this study, COVID-19 vaccination significantly increased the risks of non-serious AEs except for visual impairments (HR, 3.935; 95% CI, 0.943-16.410), with the highest level of alopecia (HR, 2.397; 95% CI, 1.896-3.029) followed by inner ear diseases (HR, 2.368; 95% CI, 2.153-2.604), and herpes zoster (HR, 2.337; 95% CI, 2.122-2.573) (Fig. 2a). In the multivariate logistic model in this study, the COVID-19 vaccination was associated with a significant increase in the risk of most non-serious AEs, indicating potential influence in the early time point (one-week after COVID-19 vaccination). At three months post-vaccination, the COVID-19 vaccination significantly increased the risk of endometriosis (OR, 1.631; 95% CI, 1.313-2.025). Furthermore, visual impairment at three months (OR, 3.956; 95% CI, 0.949-16.496), tinnitus at one-week (OR, 1.840; 95% CI, 0.981-3.451), and two weeks (OR, 1.513; 95% CI, 1.000-2.287), and periodontal diseases at one-week (OR, 0.965; 95% CI, 0.366-2.546), two weeks (OR, 1.771; 95% CI, 0.760-4.126), and one-month (OR, 1.662; 95% CI, 0.955-3.041) showed no statistical differences of ORs between two groups (Fig. 2b).

**Fig. 2.**
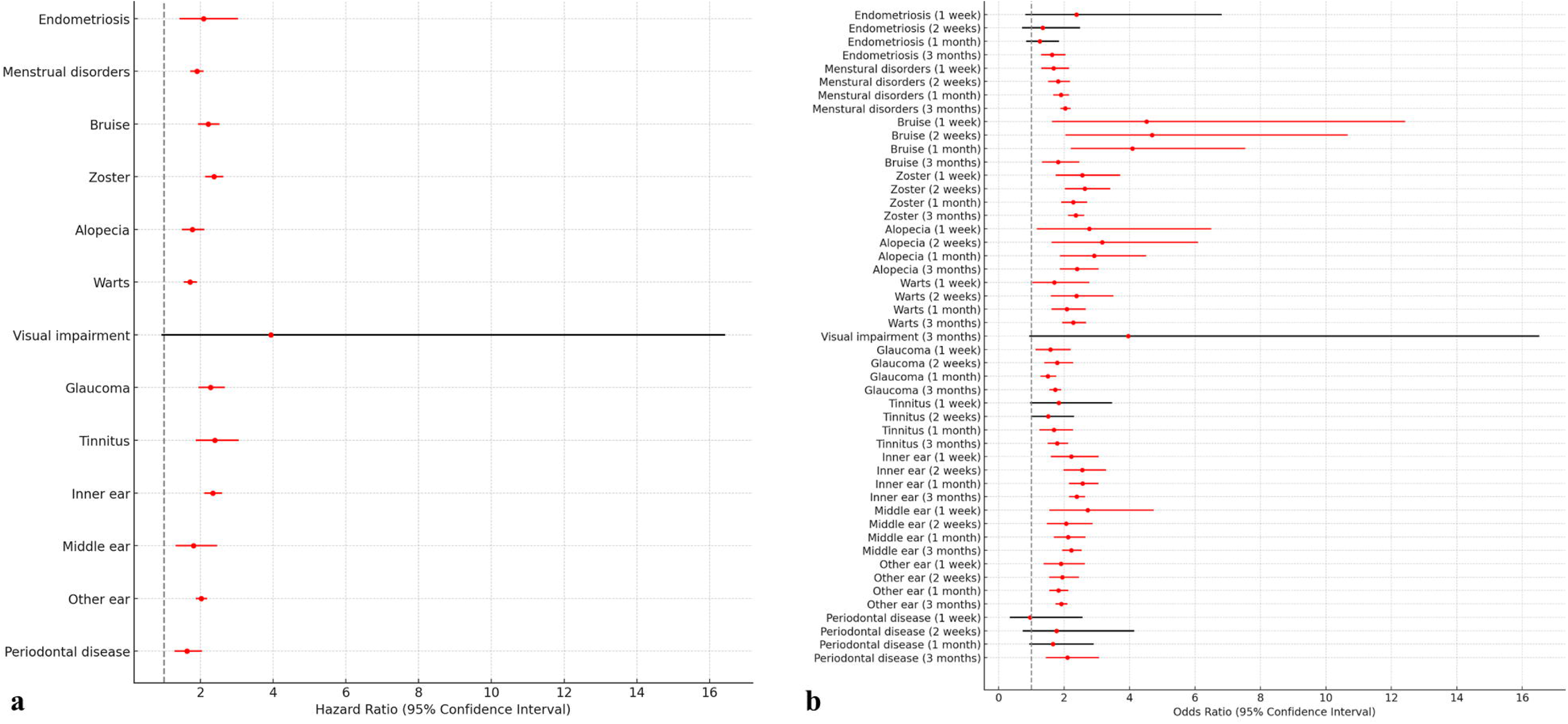
The risk for non-serious autoimmune-related adverse events (AEs) by COVID-19 vaccination. a. Cox proportional hazard model, which is presented as a forest plot (hazard ratio [red circle] with 95% confidence interval [bar]). If clinical significance, it is marked as red-bar. b. Multivariate logistic regression model along with time point, which presented as forest plot (odd ratio [red circle] with 95% confidence interval [bar]).

### The risks of non-serious AEs according to the COVID-19 vaccine type

Both the multivariate logistic regression model and the Cox proportional hazard model were used to assess the risk factors according to the COVID-19 vaccine type (Fig. 3). In the Cox proportional hazard model, heterologous vaccination was more increased the risks of gynecological problems including endometriosis (HR, 2.784; 95% CI, 2.083-3.722]) and menstrual disorders (HR, 2.837; 95% CI, 2.583-3.117]), hematological problem including bruise (HR, 1.891; 95% CI, 1.208-2.962), dermatological problems including herpes zoster (HR, 2.894; 95% CI, 2.153-2.604) and alopecia (HR, 3.413; 95% CI, 2.520-4.623), ophthalmological problem including glaucoma (HR, 1.828; 95% CI, 1.596-2.093), and periodontal diseases (HR, 3.560; 95% CI, 2.179-5.818) compared to other types of vaccination (Fig. 3a). In the multivariate logistic regression model, most trends of risks were shown a similar pattern to that of Cox proportional hazard models. According to time points, alopecia and periodontal diseases were associated with the highest risk outcomes following heterologous vaccinations compared to other vaccination methods. However, vaccination using cDNA only was observed to notably increase risks of bruise at one week (OR [95% CI] = 5.767 [2.010-16.543]), two weeks (HR, 6.260; 95% CI, 2.280-14.621), and one-month (HR, 4.996; 95% CI, 2.659-9.388]) (Fig. 3b).

**Fig. 3.**
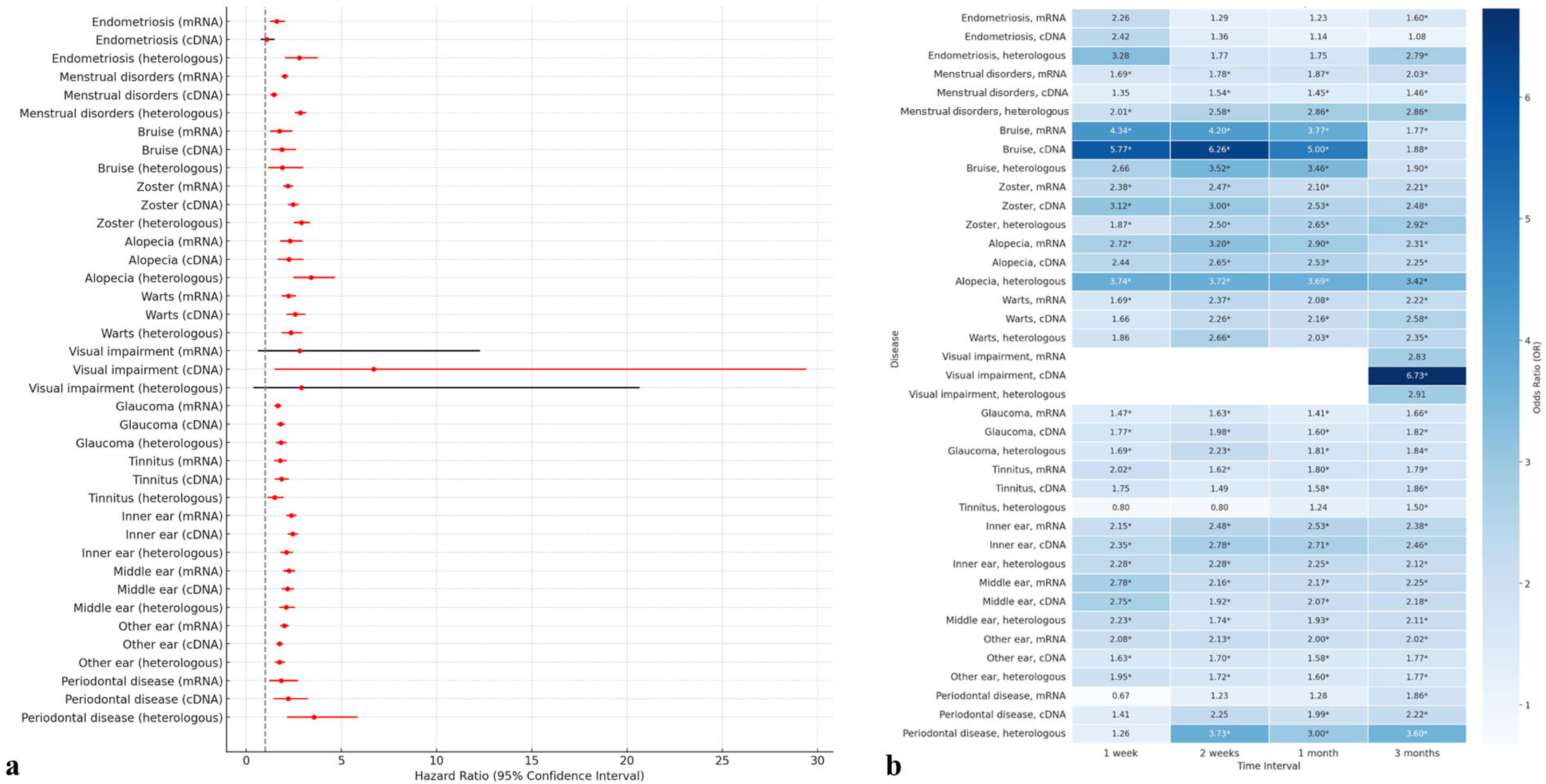
The risk for non-serious autoimmune-related adverse events (AEs) of vaccinated subjects according to vaccine type. a. Cox proportional hazard model, which is presented as a forest plot (hazard ratio [red circle] with 95% confidence interval [bar]). If clinical significance, it is marked as red-bar. b. The multivariate logistic regression model along with the time point was presented as the odd ratio of a heatmap. If clinical significance, it is marked as *.

## Discussion

The concerns for post-sequelae and vaccine-related complications have been raised as one of the global important issues [1, 9]. The main focus of the COVID-19 vaccine-related complications were serious AEs such as cardiovascular, and neurological problems that can give rise to fatal conditions [6, 7] To the best of our knowledge, the research regarding non-serious AEs following COVID-19 vaccination did not conduct a large population-based cohort study at the nationwide level [10] Here, we investigated the 14 non-serious AEs following COVID-19 vaccination in Seoul, South Korea. By comparing the 1,458,557 vaccinated subjects and 289,579 non-vaccinated subjects, we found that 13 non-serious AEs except for endometriosis showed a significant increase in cumulative incidence following COVID-19 vaccination with notable significant higher rates of menstrual disorders, herpes zoster, glaucoma, inner ear diseases, middle ear diseases, and other ear disease than in non-vaccinated subjects. Furthermore, bruise is associated with COVID-19 vaccination in the early phase showing the highest ORs at one week and two weeks. For three months follow-up, alopecia showed the highest level of HRs following COVID-19 vaccination.

The type of vaccination has been associated with immune response given cellular mechanisms [13, 23] Lee et al. showed that heterologous vaccination leads to enriched B cells and CD4^+^ T cell responses with higher activation of interferon pathways, suggesting the potential increase of ir AEs [24]. In this study for non-serious AEs, heterologous vaccination showed the highest risks of eight AEs including endometriosis, menstrual disorders, bruise, herpes zoster, alopecia, glaucoma, and periodontal diseases compared to other types of vaccination. Thus, Peripheral blood and skin lesions may exhibit heightened immune responses following heterologous vaccination.

The spike protein is considered a primary target for the development of vaccines against COVID-19 because the infection by severe acute respiratory syndrome coronavirus 2 (SARS-CoV-2) is initiated by the binding of spike protein to the ACE2 receptor on the host cell surface [23, 25]. Yonker et al. suggested that the circulating spike protein was detected in the peripheral blood of patients who developed post-mRNA vaccine myocarditis [25]. Several studies, supporting Yonker et al., provide potential insight into the possibility that mRNA-LNP can act as a potential underlying cause for diverse AEs [25–28]. The main difference between cDNA-based vaccines and mRNA-based vaccines against SARS-CoV-2 was mediators of immune responses, which were respectively spike protein and lipid-nanoparticle-encapsulated mRNA [23]. The current hypothesis between COVID-19 vaccination and ear diseases as AEs is that the ear disease is activated by intensification of a spike protein-specific IgG and potential systematic immune response suggesting the immunologic important factors [29]. Surprisingly, in this study, most ear problems including tinnitus, inner ear disease, and middle ear diseases showed the highest HRs in vaccination using cDNA only compared to other types of vaccination. The findings of this study are not only consistent with existing hypotheses indicating the important role of spike protein in SARS-CoV-2.

For gynecological and hematological issues, previous studies have suggested that the COVID-19 vaccination increased bleeding from the changes in the irregular menstrual cycle [30]. Population-based studies reported that unexpected vaginal bleeding and menstrual bleeding changes as an emerging phenomenon [27, 28]. Likewise, our study also showed that increased cIRs and risks of menstrual disorders (including menorrhagia, metrorrhagia, and hypermenorrhea), and bruises (vaccine-specific manifestation, i.e. non-tender yellow-colored bruises on especially extremities), which significantly rose on heterologous vaccinations. One of the important points for this result was the trend of the diminishing risk of bruises after vaccination. Considering previous studies, these manifestations were caused by hormonal changes resulting from spike proteins and disruption of the coagulation pathway in the endometrium [27, 28] Thus, both clinicians and vaccinated subjects need to be cautious of bruise occurrence within one-month post-vaccination. In addition, vaccination using cDNA only significantly increased the risks of bruises very significantly at the early phase compared to other types so special caution may be needed in the vaccinated subjects using cDNA vaccine up to at least two weeks post-vaccination. While there were no significant differences in cIR of endometriosis at three months between vaccinated and non-vaccinated subjects, this study revealed an increased risk of endometriosis associated with COVID-19 vaccination at the three months post-vaccination. These results suggest that the cIR of endometriosis may see a significant rise beyond three months, indicating the need for extended long-term follow-up studies.

Our study has several strengths for non-serious AEs following COVID-19 vaccination. First, many non-serious AEs have been reported as case reports or case series so it is the first study to investigate non-serious AEs following COVID-19 vaccination [14, 15, 20, 29, 31]. For diverse manifestations including post-vaccination glaucoma, ear diseases, and alopecia, these AEs shared potential pathophysiological mechanisms as expression of spike proteins, which dysregulate immunity, lead to molecular mimicry phenomena, and activate the pro-inflammatory cytokines [13, 15]. Especially, mRNA vaccine translation can be influenced by ribonucleotides as a consequence of N(1)-methylpeudouridine-induced ribosome stalling indicating a potentially pathophysiological change in human [32]. With the suggested mechanisms in the literature, our studies are consistent with this hypothesis because the types of vaccinations have higher HRs in heterologous or cDNA vaccination compared to mRNA-only vaccination. Second, the current studies have demonstrated that the COVID-19 vaccine affects T cell-mediated immune response in multiple sclerosis, which leads to autoimmunity [19]. Our findings for the spectrum of non-serious AEs strengthen their studies sharing similar pathophysiological mechanisms and hypotheses such as the role of spike proteins and autoimmune diseases triggered by vaccines. Last, the non-serious AEs after COVID-19 vaccination are relatively common and can be affected by various factors such as vaccination methods [19]. Furthermore, our findings also suggest that different types of vaccinations exhibit distinct activation patterns at various sites, which will need to be studied in the future. Even in this case, the warts on the cheek were developed with positive for spike IgG and negative for nucleocapsid IgG after mRNA-based vaccination, suggesting that the mRNA-LNP triggers an autoimmune response [18] Therefore, the COVID-19 vaccines may not be fatal, but vaccinated people with a predisposition may be more susceptible to the occurrence of AEs. Furthermore, our study indicates that it is essential to consider potential side effects that persist beyond three months. As the toxicity of COVID-19 diminishes and a significant portion of the worldwide population acquires natural immunity, it is important to designate as vaccine recipients those for whom the benefits of vaccination outweigh the potential side effects of ongoing vaccination.

This study has several limitations. Our study has statistical inequity between the two groups, which is caused by a general population-based study. However, for non-serious AEs, large-sized population study was scarce so studies through the process of propensity score matching may be necessary for future research as a new paper. Second, target AEs were extracted based on ICD-10 codes in the claim databases so coding, mismatching, or misclassification errors could have occurred. Third, we tried to broad-spectrum non-serious AEs following COVID-19 vaccination but there is a possibility that some diseases may not be included. Long-term studies are needed on the duration of AEs and on inflammatory disease-related side effects that did not appear in the short-term duration, which are underway to elucidate the AEs of COVID-19 vaccination. This should be illustrated through new research by specialty in the future.

## Conclusions

The three-month risks of incidental non-serious AEs are substantially higher in the COVID-19 vaccinated subjects than in non-vaccinated controls. Our findings suggested that vaccinated subjects with predisposition are potentially vulnerable to the occurrence of diverse AEs although the COVID-19 vaccines may not be fatal. Consequently, clinicians should maintain closed observation of a range of non-serious AEs after vaccination, given that these manifestations might emerge post-vaccination.

## Data Availability

All data produced in the present study are available upon reasonable request to the authors

## Statements and Declarations

### Funding

The authors declare that no funds, grants, or other support were received during the preparation of this manuscript.

### Competing Interests

The authors have no relevant financial or non-financial interests to disclose.

### Ethics approval and Consent to participate

This study was performed in line with the principles of the Declaration of Helsinki. Approval was granted by the Institutional Review Board of our institute (IRB No.: EUMC 2022-01-003), which waived the requirement for informed consent because data analyses were performed retrospectively using anonymized data derived from the South Korean NHIS database.

### Author contributions

JHS and HJK contributed equally as first authors. EMC conceptualized the study. JHS, M-HK, and EMC designed the study. JHS, HJK, M-HK, and EMC analysed and interpreted the data. M-HK acquired the data. JHS, HJK, and EMC drafted the manuscript. JHS, HJK, M-HK, MGC, and EMC critically reviewed the work. MGC verified the data in the study. All authors had full access to all the data and had final responsibility for the decision to submit for publication.

## Notes

### Competing Interest Statement

The authors have declared no competing interest.

### Funding Statement

This study did not receive any funding

### Author Declarations

IRB of Ewha Womans University Medical Center gave ethical approval (IRB No.: EUMC 2022-07-003) for this work.

### Summary of Updates

The title updated to clarify the diseases target

