## Supplementary Tables for "Broad-spectrum of non-serious adverse events following COVID-19 vaccination: A population-based cohort study in Seoul, South Korea"

**eTable 1. The cumulative incidence rates of non-fatal immune-related adverse events stratified by gender**

| Disease | Gender | Vaccination | total | One week | | | | Two weeks | | | | One month | | | | Three months | | | |
|---|---|---|---|---|---|---|---|---|---|---|---|---|---|---|---|---|---|---|---|
| | | | | event | IR | 95% CI | *P* | event | IR | 95% CI | *P* | event | IR | 95% CI | *P* | event | IR | 95% CI | *P* |
| Endometriosis | Male | No | 143128 | | | | | | | | | | | | | | | | |
| | | Yes | 718173 | | | | | | | | | | | | | | | | |
| | Female | No | 146448 | 4 | 0.27 | 0.01-0.54 | 0.389 | 13 | 0.89 | 0.41-1.37 | 0.877 | 35 | 2.39 | 1.60-3.18 | 0.703 | 99 | 6.76 | 5.43-8.09 | 0.164 |
| | | Yes | 740384 | 34 | 0.46 | 0.30-0.61 | | 63 | 0.85 | 0.64-1.06 | | 165 | 2.23 | 1.89-2.57 | | 584 | 7.89 | 7.25-8.53 | |
| Menstrual disorder | Male | No | 143128 | | | | | | | | | | | | | | | | |
| | | Yes | 718173 | | | | | | | | | | | | | | | | |
| | Female | No | 146448 | 82 | 5.6 | 4.39-6.81 | 0.638 | 161 | 10.99 | 9.30-12.69 | 0.112 | 346 | 23.63 | 21.14-26.11 | 0.003 | 1025 | 69.99 | 65.72-74.26 | <0.001 |
| | | Yes | 740384 | 442 | 5.97 | 5.41-6.53 | | 935 | 12.63 | 11.82-13.44 | | 2081 | 28.11 | 26.90-29.31 | | 6481 | 87.54 | 85.41-89.66 | |
| Bruise | Male | No | 143128 | 1 | 0.07 | 0.00-0.21 | 0.007 | 2 | 0.14 | 0.00-0.33 | 0.001 | 3 | 0.21 | 0.00-0.45 | <0.001 | 17 | 1.19 | 0.62-1.75 | <0.001 |
| | | Yes | 718173 | 42 | 0.58 | 0.41-0.76 | | 65 | 0.91 | 0.69-1.13 | | 108 | 1.5 | 1.22-1.79 | | 218 | 3.04 | 2.63-3.44 | |
| | Female | No | 146448 | 3 | 0.2 | 0.00-0.44 | 0.005 | 4 | 0.27 | 0.01-0.54 | <0.001 | 8 | 0.55 | 0.17-0.92 | <0.001 | 31 | 2.12 | 1.37-2.86 | <0.001 |
| | | Yes | 740384 | 65 | 0.88 | 0.66-1.09 | | 116 | 1.57 | 1.28-1.85 | | 179 | 2.42 | 2.06-2.77 | | 341 | 4.61 | 4.12-5.09 | |
| Herpes zoster | Male | No | 143128 | 8 | 0.56 | 0.17-0.95 | <0.001 | 21 | 1.47 | 0.84-2.09 | <0.001 | 59 | 4.12 | 3.07-5.17 | <0.001 | 179 | 12.51 | 10.68-14.34 | <0.001 |
| | | Yes | 718173 | 188 | 2.62 | 2.24-2.99 | | 411 | 5.72 | 5.17-6.28 | | 907 | 12.63 | 11.81-13.45 | | 2641 | 36.77 | 35.37-38.17 | |
| | Female | No | 146448 | 23 | 1.57 | 0.93-2.21 | <0.001 | 45 | 3.07 | 2.18-3.97 | <0.001 | 103 | 7.03 | 5.68-8.39 | <0.001 | 275 | 18.78 | 16.56-21.00 | <0.001 |
| | | Yes | 740384 | 284 | 3.84 | 3.39-4.28 | | 644 | 8.7 | 8.03-9.37 | | 1363 | 18.41 | 17.43-19.39 | | 3934 | 53.13 | 51.48-54.79 | |
| Alopecia | Male | No | 143128 | 2 | 0.14 | 0.00-0.33 | 0.053 | 5 | 0.35 | 0.04-0.66 | 0.047 | 11 | 0.77 | 0.31-1.22 | 0.001 | 38 | 2.65 | 1.81-3.50 | <0.001 |
| | | Yes | 718173 | 38 | 0.53 | 0.36-0.70 | | 62 | 0.86 | 0.65-1.08 | | 143 | 1.99 | 1.66-2.32 | | 373 | 5.19 | 4.67-5.72 | |
| | Female | No | 146448 | 4 | 0.27 | 0.01-0.54 | 1 | 5 | 0.34 | 0.04-0.64 | 0.064 | 12 | 0.82 | 0.36-1.28 | 0.015 | 42 | 2.87 | 2.00-3.74 | <0.001 |
| | | Yes | 740384 | 23 | 0.31 | 0.18-0.44 | | 60 | 0.81 | 0.61-1.02 | | 123 | 1.66 | 1.37-1.95 | | 393 | 5.31 | 4.78-5.83 | |
| Warts | Male | No | 143128 | 9 | 0.63 | 0.22-1.04 | 0.286 | 14 | 0.98 | 0.47-1.49 | 0.003 | 40 | 2.79 | 1.93-3.66 | <0.001 | 103 | 7.2 | 5.81-8.59 | <0.001 |
| | | Yes | 718173 | 68 | 0.95 | 0.72-1.17 | | 156 | 2.17 | 1.83-2.51 | | 349 | 4.86 | 4.35-5.37 | | 983 | 13.69 | 12.83-14.54 | |
| | Female | No | 146448 | 11 | 0.75 | 0.31-1.19 | 0.649 | 16 | 1.09 | 0.56-1.63 | 0.02 | 38 | 2.59 | 1.77-3.42 | 0.005 | 94 | 6.42 | 5.12-7.72 | <0.001 |
| | | Yes | 740384 | 67 | 0.9 | 0.69-1.12 | | 147 | 1.99 | 1.66-2.31 | | 309 | 4.17 | 3.71-4.64 | | 828 | 11.18 | 10.42-11.94 | |
| Visual impairment | Male | No | 143128 | 0 | 0 | 0.00-0.00 | 1 | 0 | 0 | 0.00-0.00 | 0.598 | 0 | 0 | 0.00-0.00 | 0.154 | 0 | 0 | 0.00-0.00 | 0.006 |
| | | Yes | 718173 | 3 | 0.04 | 0.00-0.09 | | 6 | 0.08 | 0.02-0.15 | | 15 | 0.21 | 0.10-0.31 | | 31 | 0.43 | 0.28-0.58 | |
| | Female | No | 146448 | 0 | 0 | 0.00-0.00 | 1 | 0 | 0 | 0.00-0.00 | 1 | 0 | 0 | 0.00-0.00 | 0.368 | 2 | 0.14 | 0.00-0.33 | 0.561 |
| | | Yes | 740384 | 2 | 0.03 | 0.00-0.06 | | 3 | 0.04 | 0.00-0.09 | | 8 | 0.11 | 0.03-0.18 | | 19 | 0.26 | 0.14-0.37 | |
| Glaucoma | Male | No | 143128 | 11 | 0.77 | 0.31-1.22 | <0.001 | 28 | 1.96 | 1.23-2.68 | <0.001 | 78 | 5.45 | 4.24-6.66 | <0.001 | 216 | 15.09 | 13.08-17.10 | <0.001 |
| | | Yes | 718173 | 198 | 2.76 | 2.37-3.14 | | 393 | 5.47 | 4.93-6.01 | | 849 | 11.82 | 11.03-12.62 | | 2556 | 35.59 | 34.21-36.97 | |
| | Female | No | 146448 | 32 | 2.19 | 1.43-2.94 | 0.056 | 52 | 3.55 | 2.59-4.52 | <0.001 | 121 | 8.26 | 6.79-9.73 | <0.001 | 318 | 21.71 | 19.33-24.10 | <0.001 |
| | | Yes | 740384 | 232 | 3.13 | 2.73-3.54 | | 508 | 6.86 | 6.26-7.46 | | 1043 | 14.09 | 13.23-14.94 | | 3193 | 43.13 | 41.63-44.62 | |
| Tinnitus | Male | No | 143128 | 5 | 0.35 | 0.04-0.66 | 0.197 | 12 | 0.84 | 0.36-1.31 | 0.081 | 25 | 1.75 | 1.06-2.43 | 0.001 | 73 | 5.1 | 3.93-6.27 | <0.001 |
| | | Yes | 718173 | 48 | 0.67 | 0.48-0.86 | | 105 | 1.46 | 1.18-1.74 | | 238 | 3.31 | 2.89-3.73 | | 789 | 10.99 | 10.22-11.75 | |
| | Female | No | 146448 | 6 | 0.41 | 0.08-0.74 | 0.044 | 14 | 0.96 | 0.46-1.46 | 0.025 | 29 | 1.98 | 1.26-2.70 | <0.001 | 98 | 6.69 | 5.37-8.02 | <0.001 |
| | | Yes | 740384 | 71 | 0.96 | 0.74-1.18 | | 132 | 1.78 | 1.48-2.09 | | 296 | 4 | 3.54-4.45 | | 1000 | 13.51 | 12.67-14.34 | |
| Inner ear disease | Male | No | 143128 | 13 | 0.91 | 0.41-1.40 | <0.001 | 20 | 1.4 | 0.78-2.01 | <0.001 | 48 | 3.35 | 2.41-4.30 | <0.001 | 147 | 10.27 | 8.61-11.93 | <0.001 |
| | | Yes | 718173 | 171 | 2.38 | 2.02-2.74 | | 355 | 4.94 | 4.43-5.46 | | 798 | 11.11 | 10.34-11.88 | | 2219 | 30.9 | 29.61-32.18 | |
| | Female | No | 146448 | 30 | 2.05 | 1.32-2.78 | <0.001 | 51 | 3.48 | 2.53-4.44 | <0.001 | 104 | 7.1 | 5.74-8.47 | <0.001 | 319 | 21.78 | 19.39-24.17 | <0.001 |
| | | Yes | 740384 | 382 | 5.16 | 4.64-5.68 | | 739 | 9.98 | 9.26-10.70 | | 1583 | 21.38 | 20.33-22.43 | | 4651 | 62.82 | 61.02-64.62 | |
| Middle ear disease | Male | No | 143128 | 5 | 0.35 | 0.04-0.66 | <0.001 | 15 | 1.05 | 0.52-1.58 | <0.001 | 39 | 2.72 | 1.87-3.58 | <0.001 | 126 | 8.8 | 7.27-10.34 | <0.001 |
| | | Yes | 718173 | 102 | 1.42 | 1.14-1.70 | | 217 | 3.02 | 2.62-3.42 | | 495 | 6.89 | 6.29-7.50 | | 1495 | 20.82 | 19.76-21.87 | |
| | Female | No | 146448 | 9 | 0.61 | 0.21-1.02 | 0.004 | 27 | 1.84 | 1.15-2.54 | 0.002 | 54 | 3.69 | 2.70-4.67 | <0.001 | 164 | 11.2 | 9.49-12.91 | <0.001 |
| | | Yes | 740384 | 116 | 1.57 | 1.28-1.85 | | 251 | 3.39 | 2.97-3.81 | | 563 | 7.6 | 6.98-8.23 | | 1848 | 24.96 | 23.82-26.10 | |
| Other ear disease | Male | No | 143128 | 19 | 1.33 | 0.73-1.92 | <0.001 | 32 | 2.24 | 1.46-3.01 | <0.001 | 84 | 5.87 | 4.61-7.12 | <0.001 | 267 | 18.65 | 16.42-20.89 | <0.001 |
| | | Yes | 718173 | 260 | 3.62 | 3.18-4.06 | | 526 | 7.32 | 6.70-7.95 | | 1134 | 15.79 | 14.87-16.71 | | 3460 | 48.18 | 46.58-49.78 | |
| | Female | No | 146448 | 24 | 1.64 | 0.98-2.29 | <0.001 | 54 | 3.69 | 2.70-4.67 | <0.001 | 118 | 8.06 | 6.60-9.51 | <0.001 | 340 | 23.22 | 20.75-25.68 | <0.001 |
| | | Yes | 740384 | 290 | 3.92 | 3.47-4.37 | | 586 | 7.91 | 7.27-8.56 | | 1307 | 17.65 | 16.70-18.61 | | 4092 | 55.27 | 53.58-56.96 | |
| Periodontal disease | Male | No | 143128 | 0 | 0 | 0.00-0.00 | 0.037 | 0 | 0 | 0.00-0.00 | 0.003 | 5 | 0.35 | 0.04-0.66 | 0.01 | 11 | 0.77 | 0.31-1.22 | <0.001 |
| | | Yes | 718173 | 21 | 0.29 | 0.17-0.42 | | 36 | 0.5 | 0.34-0.67 | | 75 | 1.04 | 0.81-1.28 | | 247 | 3.44 | 3.01-3.87 | |
| | Female | No | 146448 | 5 | 0.34 | 0.04-0.64 | 0.067 | 6 | 0.41 | 0.08-0.74 | 1 | 9 | 0.61 | 0.21-1.02 | 0.071 | 20 | 1.37 | 0.77-1.96 | <0.001 |
| | | Yes | 740384 | 9 | 0.12 | 0.04-0.20 | | 34 | 0.46 | 0.30-0.61 | | 85 | 1.15 | 0.90-1.39 | | 241 | 3.26 | 2.84-3.67 | |

**eTable 2. The cumulative incidence rates of non-fatal immune-related adverse events stratified by vaccine type**

| Disease | Vaccination | total | One week | | | | Two weeks | | | | One month | | | | Three months | | | |
|---|---|---|---|---|---|---|---|---|---|---|---|---|---|---|---|---|---|---|
| | | | event | IR | 95% CI | *P* | event | IR | 95% CI | *P* | event | IR | 95% CI | *P* | event | IR | 95% CI | *P* |
| Endometriosis | No | 289576 | 4 | 0.14 | 0.00-0.27 | 0.264 | 13 | 0.45 | 0.20-0.69 | 0.213 | 35 | 1.21 | 0.81-1.61 | 0.002 | 99 | 3.42 | 2.75-4.09 | <0.001 |
| | only mRNA vaccine | 849526 | 22 | 0.26 | 0.15-0.37 | | 41 | 0.48 | 0.33-0.63 | | 109 | 1.28 | 1.04-1.52 | | 401 | 4.72 | 4.26-5.18 | |
| | only cDNA vaccine | 510253 | 8 | 0.16 | 0.05-0.27 | | 15 | 0.29 | 0.15-0.44 | | 37 | 0.73 | 0.49-0.96 | | 97 | 1.9 | 1.52-2.28 | |
| | Cross | 98778 | 4 | 0.4 | 0.01-0.80 | | 7 | 0.71 | 0.18-1.23 | | 19 | 1.92 | 1.06-2.79 | | 86 | 8.71 | 6.87-10.55 | |
| Menstrual disorder | No | 289576 | 82 | 2.83 | 2.22-3.44 | <0.001 | 161 | 5.56 | 4.70-6.42 | <0.001 | 346 | 11.95 | 10.69-13.21 | <0.001 | 1025 | 35.4 | 33.23-37.56 | <0.001 |
| | only mRNA vaccine | 849526 | 334 | 3.93 | 3.51-4.35 | | 685 | 8.06 | 7.46-8.67 | | 1533 | 18.05 | 17.14-18.95 | | 4891 | 57.57 | 55.96-59.18 | |
| | only cDNA vaccine | 510253 | 63 | 1.23 | 0.93-1.54 | | 138 | 2.7 | 2.25-3.16 | | 281 | 5.51 | 4.86-6.15 | | 810 | 15.87 | 14.78-16.97 | |
| | Cross | 98778 | 45 | 4.56 | 3.22-5.89 | | 112 | 11.34 | 9.24-13.44 | | 267 | 27.03 | 23.79-30.27 | | 780 | 78.96 | 73.45-84.48 | |
| Bruise | No | 289576 | 4 | 0.14 | 0.00-0.27 | <0.001 | 6 | 0.21 | 0.04-0.37 | <0.001 | 11 | 0.38 | 0.16-0.60 | <0.001 | 48 | 1.66 | 1.19-2.13 | <0.001 |
| | only mRNA vaccine | 849526 | 56 | 0.66 | 0.49-0.83 | | 86 | 1.01 | 0.80-1.23 | | 142 | 1.67 | 1.40-1.95 | | 296 | 3.48 | 3.09-3.88 | |
| | only cDNA vaccine | 510253 | 47 | 0.92 | 0.66-1.18 | | 87 | 1.71 | 1.35-2.06 | | 131 | 2.57 | 2.13-3.01 | | 231 | 4.53 | 3.94-5.11 | |
| | Cross | 98778 | 4 | 0.4 | 0.01-0.80 | | 8 | 0.81 | 0.25-1.37 | | 14 | 1.42 | 0.67-2.16 | | 32 | 3.24 | 2.12-4.36 | |
| Herpes zoster | No | 289576 | 31 | 1.07 | 0.69-1.45 | <0.001 | 66 | 2.28 | 1.73-2.83 | <0.001 | 162 | 5.59 | 4.73-6.46 | <0.001 | 454 | 15.68 | 14.24-17.12 | <0.001 |
| | only mRNA vaccine | 849526 | 237 | 2.79 | 2.43-3.14 | | 535 | 6.3 | 5.76-6.83 | | 1136 | 13.37 | 12.60-14.15 | | 3365 | 39.61 | 38.27-40.95 | |
| | only cDNA vaccine | 510253 | 215 | 4.21 | 3.65-4.78 | | 463 | 9.07 | 8.25-9.90 | | 987 | 19.34 | 18.14-20.55 | | 2760 | 54.09 | 52.08-56.10 | |
| | Cross | 98778 | 20 | 2.02 | 1.14-2.91 | | 57 | 5.77 | 4.27-7.27 | | 147 | 14.88 | 12.48-17.29 | | 450 | 45.56 | 41.36-49.76 | |
| Alopecia | No | 289576 | 6 | 0.21 | 0.04-0.37 | 0.024 | 10 | 0.35 | 0.13-0.56 | 0.001 | 23 | 0.79 | 0.47-1.12 | <0.001 | 80 | 2.76 | 2.16-3.37 | <0.001 |
| | only mRNA vaccine | 849526 | 41 | 0.48 | 0.33-0.63 | | 82 | 0.97 | 0.76-1.17 | | 175 | 2.06 | 1.75-2.37 | | 482 | 5.67 | 5.17-6.18 | |
| | only cDNA vaccine | 510253 | 13 | 0.25 | 0.12-0.39 | | 28 | 0.55 | 0.35-0.75 | | 63 | 1.23 | 0.93-1.54 | | 195 | 3.82 | 3.29-4.36 | |
| | Cross | 98778 | 7 | 0.71 | 0.18-1.23 | | 12 | 1.21 | 0.53-1.90 | | 28 | 2.83 | 1.78-3.88 | | 89 | 9.01 | 7.14-10.88 | |
| Warts | No | 289576 | 20 | 0.69 | 0.39-0.99 | 0.052 | 30 | 1.04 | 0.67-1.41 | <0.001 | 78 | 2.69 | 2.10-3.29 | <0.001 | 197 | 6.8 | 5.85-7.75 | <0.001 |
| | only mRNA vaccine | 849526 | 89 | 1.05 | 0.83-1.27 | | 197 | 2.32 | 2.00-2.64 | | 433 | 5.1 | 4.62-5.58 | | 1157 | 13.62 | 12.84-14.40 | |
| | only cDNA vaccine | 510253 | 34 | 0.67 | 0.44-0.89 | | 79 | 1.55 | 1.21-1.89 | | 173 | 3.39 | 2.89-3.90 | | 504 | 9.88 | 9.02-10.74 | |
| | Cross | 98778 | 12 | 1.21 | 0.53-1.90 | | 27 | 2.73 | 1.70-3.76 | | 52 | 5.26 | 3.83-6.69 | | 150 | 15.19 | 12.76-17.61 | |
| Visual impairment | No | 289576 | 0 | 0 | 0.00-0.00 | 0.421 | 0 | 0 | 0.00-0.00 | 0.08 | 0 | 0 | 0.00-0.00 | 0.012 | 2 | 0.07 | 0.00-0.16 | <0.001 |
| | only mRNA vaccine | 849526 | 2 | 0.02 | 0.00-0.06 | | 3 | 0.04 | 0.00-0.08 | | 8 | 0.09 | 0.03-0.16 | | 18 | 0.21 | 0.11-0.31 | |
| | only cDNA vaccine | 510253 | 3 | 0.06 | 0.00-0.13 | | 6 | 0.12 | 0.02-0.21 | | 13 | 0.25 | 0.12-0.39 | | 30 | 0.59 | 0.38-0.80 | |
| | Cross | 98778 | 0 | 0 | 0.00-0.00 | | 0 | 0 | 0.00-0.00 | | 2 | 0.2 | 0.00-0.48 | | 2 | 0.2 | 0.00-0.48 | |
| Glaucoma | No | 289576 | 43 | 1.48 | 1.04-1.93 | <0.001 | 80 | 2.76 | 2.16-3.37 | <0.001 | 199 | 6.87 | 5.92-7.83 | <0.001 | 534 | 18.44 | 16.88-20.00 | <0.001 |
| | only mRNA vaccine | 849526 | 218 | 2.57 | 2.23-2.91 | | 449 | 5.29 | 4.80-5.77 | | 966 | 11.37 | 10.65-12.09 | | 3034 | 35.71 | 34.45-36.98 | |
| | only cDNA vaccine | 510253 | 186 | 3.65 | 3.12-4.17 | | 389 | 7.62 | 6.87-8.38 | | 798 | 15.64 | 14.56-16.72 | | 2367 | 46.39 | 44.52-48.25 | |
| | Cross | 98778 | 26 | 2.63 | 1.62-3.64 | | 63 | 6.38 | 4.80-7.95 | | 128 | 12.96 | 10.71-15.20 | | 348 | 35.23 | 31.54-38.93 | |
| Tinnitus | No | 289576 | 11 | 0.38 | 0.16-0.60 | 0.019 | 26 | 0.9 | 0.55-1.24 | 0.002 | 54 | 1.86 | 1.37-2.36 | <0.001 | 171 | 5.91 | 5.02-6.79 | <0.001 |
| | only mRNA vaccine | 849526 | 72 | 0.85 | 0.65-1.04 | | 138 | 1.62 | 1.35-1.90 | | 316 | 3.72 | 3.31-4.13 | | 991 | 11.67 | 10.94-12.39 | |
| | only cDNA vaccine | 510253 | 44 | 0.86 | 0.61-1.12 | | 92 | 1.8 | 1.43-2.17 | | 195 | 3.82 | 3.29-4.36 | | 708 | 13.88 | 12.85-14.90 | |
| | Cross | 98778 | 3 | 0.3 | 0.00-0.65 | | 7 | 0.71 | 0.18-1.23 | | 23 | 2.33 | 1.38-3.28 | | 90 | 9.11 | 7.23-10.99 | |
| Inner ear disease | No | 289576 | 43 | 1.48 | 1.04-1.93 | <0.001 | 71 | 2.45 | 1.88-3.02 | <0.001 | 152 | 5.25 | 4.41-6.08 | <0.001 | 466 | 16.09 | 14.63-17.55 | <0.001 |
| | only mRNA vaccine | 849526 | 297 | 3.5 | 3.10-3.89 | | 582 | 6.85 | 6.29-7.41 | | 1285 | 15.13 | 14.30-15.95 | | 3735 | 43.97 | 42.56-45.37 | |
| | only cDNA vaccine | 510253 | 222 | 4.35 | 3.78-4.92 | | 456 | 8.94 | 8.12-9.76 | | 977 | 19.15 | 17.95-20.35 | | 2793 | 54.74 | 52.71-56.76 | |
| | Cross | 98778 | 34 | 3.44 | 2.29-4.60 | | 56 | 5.67 | 4.18-7.15 | | 119 | 12.05 | 9.88-14.21 | | 342 | 34.62 | 30.96-38.29 | |
| Middle ear disease | No | 289576 | 14 | 0.48 | 0.23-0.74 | <0.001 | 42 | 1.45 | 1.01-1.89 | <0.001 | 93 | 3.21 | 2.56-3.86 | <0.001 | 290 | 10.01 | 8.86-11.17 | <0.001 |
| | only mRNA vaccine | 849526 | 125 | 1.47 | 1.21-1.73 | | 284 | 3.34 | 2.95-3.73 | | 623 | 7.33 | 6.76-7.91 | | 1973 | 23.22 | 22.20-24.25 | |
| | only cDNA vaccine | 510253 | 82 | 1.61 | 1.26-1.95 | | 158 | 3.1 | 2.61-3.58 | | 372 | 7.29 | 6.55-8.03 | | 1157 | 22.68 | 21.37-23.98 | |
| | Cross | 98778 | 11 | 1.11 | 0.46-1.77 | | 26 | 2.63 | 1.62-3.64 | | 63 | 6.38 | 4.80-7.95 | | 213 | 21.56 | 18.67-24.46 | |
| Other ear disease | No | 289576 | 43 | 1.48 | 1.04-1.93 | <0.001 | 86 | 2.97 | 2.34-3.60 | <0.001 | 202 | 6.98 | 6.01-7.94 | <0.001 | 607 | 20.96 | 19.30-22.63 | <0.001 |
| | only mRNA vaccine | 849526 | 323 | 3.8 | 3.39-4.22 | | 655 | 7.71 | 7.12-8.30 | | 1448 | 17.04 | 16.17-17.92 | | 4309 | 50.72 | 49.21-52.23 | |
| | only cDNA vaccine | 510253 | 200 | 3.92 | 3.38-4.46 | | 408 | 8 | 7.22-8.77 | | 886 | 17.36 | 16.22-18.51 | | 2881 | 56.46 | 54.41-58.52 | |
| | Cross | 98778 | 27 | 2.73 | 1.70-3.76 | | 49 | 4.96 | 3.57-6.35 | | 107 | 10.83 | 8.78-12.88 | | 362 | 36.65 | 32.88-40.42 | |
| Periodontal disease | No | 289576 | 5 | 0.17 | 0.02-0.32 | 0.081 | 6 | 0.21 | 0.04-0.37 | <0.001 | 14 | 0.48 | 0.23-0.74 | <0.001 | 31 | 1.07 | 0.69-1.45 | <0.001 |
| | only mRNA vaccine | 849526 | 11 | 0.13 | 0.05-0.21 | | 26 | 0.31 | 0.19-0.42 | | 65 | 0.77 | 0.58-0.95 | | 226 | 2.66 | 2.31-3.01 | |
| | only cDNA vaccine | 510253 | 17 | 0.33 | 0.17-0.49 | | 37 | 0.73 | 0.49-0.96 | | 82 | 1.61 | 1.26-1.95 | | 228 | 4.47 | 3.89-5.05 | |
| | Cross | 98778 | 2 | 0.2 | 0.00-0.48 | | 7 | 0.71 | 0.18-1.23 | | 13 | 1.32 | 0.60-2.03 | | 34 | 3.44 | 2.29-4.60 | |
